# Center Related Variation in Hospitalization Cost for Patients undergoing Percutaneous Left Atrial Appendage Occlusion

**DOI:** 10.1101/2023.05.22.23290370

**Authors:** Shivaraj Patil, Chaitanya Rojulpote, Abhijit Bhattaru, Avica Atri, Viha Atri, Ola Khraisha, Sumeet Mainigi

## Abstract

**Background:** The commercial use of percutaneous LAAO with the Watchman device is increasing in the United States. The purpose of this study was to evaluate center related variation in total hospital costs for Watchman device implantation and identify factors associated with high hospital costs at a national level.

**Methods:** All adults undergoing elective LAAO with Watchman were identified in the 2016 to 2018 National Inpatient Database. Mixed models were used to evaluate the impact of center on total hospital costs adjusting for patient and center characteristics, and length of stay.

**Results:** A total of 30,175 patients underwent Watchman device implantation at a median cost of $24,500 and demonstrated significant variability across admissions (inter-decile range, $13,900-37,000). Nearly 13% of the variability in patient-level costs was related to the center performing the procedure rather than patient factors. Higher volume centers had lower total costs and demonstrated lesser total cost variation. Centers with low procedural volume, occurrence of procedural complications, congestive heart failure, and length of stay were independent predictors of a high-cost hospitalization. Though complications were associated with increased expenditure, they did not explain the observed cost variation related to the center.

**Conclusion:** A significant proportion of variation in total hospital cost was attributable to the center performing the procedure. Addressing variability of Watchman-related costs is necessary to achieve high-quality value-based care.

## Introduction

Stroke among atrial fibrillation (AF) patients is a major cause of disability and substantial economic burden^1^. Long-term oral anticoagulation (OAC) has been the mainstay of treatment for prevention of cardioembolic stroke in AF. Percutaneous left atrial appendage occlusion (LAAO) has become an attractive alternative to reduce the risk of stroke in patients with nonvalvular AF when OAC is not suitable or hazardous^2^.

LAAO with the Watchman device (Boston Scientific, Marlborough, Massachusetts) was approved in March 2015 in the United States and is being increasingly utilized for mitigation of thromboembolic risk^3^. While the initial cost of LAAO with Watchman device is high, its proven to be a cost-effective treatment strategy compared to vitamin K antagonists (VKA) and non– vitamin K antagonist oral anticoagulant (NOAC) therapies^4,5^. With the sustained rise in US healthcare expenditures and increasing emphasis on value-based health care delivery, examination of costs associated with Watchman device implantation is particularly relevant^6^.

To date, variation in Watchman procedure related expenditures have not been studied. Thus, the purpose of this study was to evaluate center related variation in total hospital costs for Watchman device implantation and identify factors associated with high hospital costs at a national level.

## Methods

### Data Source

We performed a 3-year population-based retrospective cross-sectional analysis using national (United States) data from the January 2016 to December 2018. National Inpatient Sample (NIS) database is the largest publicly available all-payer inpatient care database from the United States. It is developed as a part of the Healthcare Cost and Utilization Project (HCUP) and is sponsored by the Agency for Healthcare Research and Quality available at https://www.hcup-us.ahrq.gov/overview.jsp. The NIS includes data from all non-federal, shortterm, general, and other specialty hospitals in the United States (excluding rehabilitation and long-term acute care hospitals) in the form of de-identified patient information containing demographics, discharge diagnoses, co-morbidities, procedures, outcomes, and hospitalization costs. All states that participate in HCUP provide data to the NIS, covering >95% of the U.S. population. The database was designed to include data from a 20% sample of discharges from all participating hospitals. This design of the NIS reduces the margin of error for estimates and delivers more stable and precise estimations. The study was exempt from an Institutional Review Board approval because HCUP-NIS is a publicly available database containing only deidentified patient information.

### Study Population

All adults (age≥18 years) who underwent elective LAAO with Watchman device were identified using International Classification of Diseases – 10th Revision (ICD-10) procedure code 02L73DK. Patients with missing data on age, sex, hospitalization costs and in-hospital mortality were excluded. Furthermore, to reduce the possibility of data duplication, patients with an indicator for transfer to another acute-care facility were excluded.

### Variable Definitions

Baseline patient characteristics including age, sex, race, income level, and payer status were defined in accordance with the NIS data dictionary. The previously validated Charlson Comorbidity Index (CCI) was used to quantify the burden of chronic conditions. In-hospital major adverse events (MAE) was defined as the composite of mortality, stroke [ischemic or hemorrhagic] or TIA, bleeding or transfusion, vascular complications, myocardial infarction, systemic embolization, and pericardial effusion or tamponade requiring pericardiocentesis or surgery. The ICD-10 codes used to define these variables are listed in **Supplemental Table 1**. Annual hospital volume was calculated as the total number of elective Watchman device implantation performed at each center. Hospitals were subsequently classified into low volume: ≤ 15 procedures/year (LVH), medium volume: 16-35 procedures/year (MVH), and high volume: ≥ 36 procedures/year (HVH) based on their annual case load. Hospitalization costs were generated by application of hospital specific cost-to-charge ratio and inflation adjusted to 2018. Total Hospitalization Costs represent the expenses incurred in the production of hospital services, such as wages, supplies, and utility. However, physician professional fees are not captured by the NIS database. Admission was designated as a high-cost hospitalization if total unadjusted hospitalization cost was in the highest decile.

### Outcome

The primary outcome was total hospitalization cost at patient-level and its variation related to center-level differences. We also analyzed the variation in MAE attributable to center level differences due to high correlation between the incidence of complications and hospitalization costs at the patient level. Secondarily, we assessed patient characteristics and predictors of high-cost hospitalization for LAAO with Watchman device.

### Statistical analysis

National estimates were calculated by applying discharge weights. Categorical variables are reported as proportions and compared using Pearson’s Chi-squared test. Continuous variables are reported as means with standard deviation (SD) or median with interquartile range (IQR), when appropriate. Means and Medians were compared using independent samples t-test and MannWhitney-U test, respectively. Median Costs from 2016 to 2018, and between LVH, MVH, HVH were compared using non-parametric, independent samples Kruskal-Wallis Test. A multivariate regression model with high-cost hospitalization status as dependent variable was developed to examine predictive factors. To evaluate the effect of individual center on total hospital costs, a 2-level generalized mixed effects model was developed with center as a random effect and adjusted for various other factors (age, female sex, white race, income quartile, payer status, MAE, comorbidities, CCI score, hospital region, hospital procedural volume and length of stay) as fixed effects. Total cost was log-transformed for analysis because of the skewed distribution of cost data. The proportion of total cost variation explained by the random center effect was calculated. SPSS Statistics 25.0 (IBM Corp., Armonk, New York) and R statistical software (R Core Team 2020) were used to perform the statistical analysis. All p values were 2-sided with a significance threshold of <0.05.

## Results

A total of 30,175 patients met the study criteria and underwent elective admission for Watchman device implantation at an average of 290 hospitals per year across the United States. The mean age was 76 years, and women constituted 41.7% of the cohort. Less than 20% of the patients were in the highest income quartile, and Medicare was the primary insurer for most patients (89%). Congestive heart failure was the most common comorbidity, and median CCI score was 1 [1-3]. The vast majority of patients (62.5%) underwent Watchman device implantation at a high-volume hospital. A major adverse event (MAE) occurred in 4.6% of the study cohort with bleeding/transfusion (2.9%) and vascular complication (2.5%) being the most common events.

In-hospital mortality was 0.1% (**Table 1**). The rates of MAEs were lower in HVH compared to LVH (4.3% vs. 5.1%, p=0.016).

**Table 1.** Baseline Characteristics

| Variable | Count | Summary Statistic |
| --- | --- | --- |
| Age (years, mean +/-SD) | - | 76.02 +/- 7.97 |
| Female (%) | 12,585 | 41.7 |
| White (%) | 25,375 | 84.1 |
| Charlson Comorbidity Index (median, IQR) | - | 1 [1-3] |
| Comorbidities (%) |  |  |
| Congestive Heart Failure | 11595 | 38.4 |
| Coronary Artery Disease | 3755 | 12.4 |
| Peripheral Vascular Disease | 4930 | 16.3 |
| Cerebrovascular Disease | 2275 | 7.5 |
| Chronic Obstructive Pulmonary Disease | 6605 | 21.9 |
| Chronic Kidney Disease | 7220 | 23.9 |
| Moderate-Severe Liver Disease | 155 | 0.5 |
| Diabetes | 10455 | 34.6 |
| Income Quartile |  |  |
| 76<sup>th</sup>-100<sup>th</sup> | 7585 | 19.8 |
| 51<sup>st</sup>-75<sup>th</sup> | 8390 | 25.8 |
| 26<sup>th</sup>-50<sup>th</sup> | 7780 | 27.8 |
| 1<sup>st</sup>-25<sup>th</sup> | 5980 | 25.1 |
| Payer Status |  |  |
| Medicare | 26850 | 89.0 |
| Medicaid | 340 | 1.1 |
| Private | 2370 | 7.9 |
| Other | 545 | 1.8 |
| Hospital Region |  |  |
| New England | 820 | 2.7 |
| Middle Atlantic | 3720 | 12.3 |
| East North Central | 4255 | 14.1 |
| West North Central | 2475 | 8.2 |
| South Atlantic | 6345 | 21 |
| East South Central | 1725 | 5.7 |
| West South Central | 3660 | 12.1 |
| Mountain | 3025 | 10 |
| Pacific | 4150 | 13.8 |
| Hospital Volume |  |  |
| Low Volume (1-15 procedures/year) | 3360 | 11.1 |
| Medium Volume (16-35 procedures/year) | 7955 | 26.4 |
| High Volume (> 35 procedures/year) | 18860 | 62.5 |
| Major Adverse Event (%) | 1385 | 4.6 |
| In-Hospital Death | 40 | 0.1 |
| Acute MI | 20 | 0.1 |
| Pericardial Effusion/Tamponade requiring pericardiocentesis or surgery | 285 | 0.9 |
| Bleeding or Transfusion | 885 | 2.9 |
| Stroke (Ischemic/Hemorrhagic) or TIA | 175 | 0.6 |
| Systemic Embolization | 30 | 0.1 |
| Vascular Complication | 755 | 2.5 |
| Median Length of Stay (days) |  | 1 |
| Median Cost (interdecile range) | | \$24,500 (\$13,900-37,000) |

On a national level, the median unadjusted patient level hospitalization cost for Watchman device implantation was $24,500 and demonstrated significant variability across admissions in our study cohort (inter-decile range, $13,900-37,000). The median hospitalization costs decreased slightly over the study period from $ 24,600 [IQR, $18,900 – 30,900] in 2016 to $ 24,400 [IQR, $18,600 – 29,800] in 2018 (p <0.001). As expected, patient-level costs were significantly (p<0.001) greater for patients experiencing a MAE: $28,700 (IQR, $21,700-37,100) compared to those who did not experience a MAE: $24,400 (IQR, $18,600-30,100). Median hospitalization costs were significantly lower in HVH: $24,000 (IQR, $18,700 – 29,200), compared to LVH: $25,900 (IQR, $20,100 – 33,400) in our study sample (p<0.001). Additionally, there was significant variation (p<0.001) in median hospitalization costs based on primary payer: Medicare: $24,600 (IQR, $19,000 – 30,400), Medicaid: $24,900 (IQR, $19,800 – 30,900), Private Pay: $24,900 (IQR, $18,000 - 30,400), Other Pay: $17,100 (IQR, $8,800 – 27,900).

Analysis of random intercept from the mixed model revealed 12.9 % of total cost variation for Watchman device implantation was due to the center-level differences. Whereas only 0.8% of the variation in complications was attributable to center-level differences. Among patients who experienced a MAE, 14.1% of inter-hospital variation in total costs was attributable to center level differences, compared to 13.2% in those who did not experience a MAE. On examining the relationship between annual hospital volume for Watchman device implantation and cost variation, we observed a decline in the proportion of cost variation attributable to the center with 14.6% in LVH and 10.9% in HVH. For patients primarily insured by Medicare, 13.3% of inter-hospital variation of total cost was attributable to center level differences, while this variation was 21.2% among non-Medicare (Medicaid, Private Pay, Other) patients (**Figure 1**).

**Figure 1.**
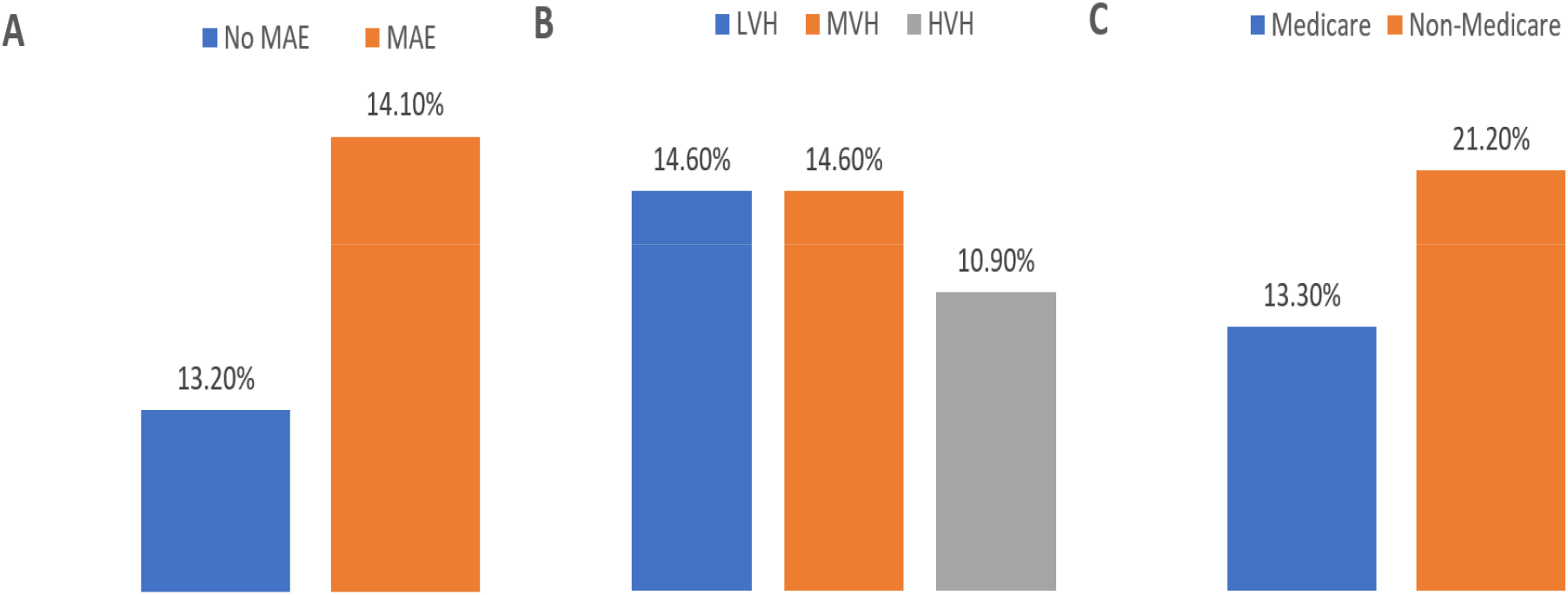
Degree of Inter-hospital Variation in cost following Watchman Implantation. Panel A shows the degree of variation among patients with and without major adverse events. Panel B shows the degree of variation based on hospital Watchman implantation volume. Panel C shows degree of variation in Medicare and Non-Medicare beneficiaries.

Admissions with total unadjusted hospitalization costs > 90th percentile ($37,000) were categorized as high-cost hospitalizations. Compared with others, patients who experienced a high-cost hospitalization had similar distribution of age and primary payer status (Medicare). High-cost hospitalization patients were more commonly women, and approximately 30% belonged to the highest income quartile. The burden of congestive heart failure and MAEs were significantly greater among high-cost hospitalizations. Amongst patients who did not experience a high-cost hospitalization, nearly 64% patients underwent Watchman device implantation at a HVH, and only 11% of patients were treated at a LVH. Whereas, patients who experienced a high-cost hospitalization, a greater proportion of patients were treated at a LVH (15.9%) and only 52% were treated at a HVH (**Table 2**).

**Table 2.** Comparison of baseline characteristics between high-cost and non-high-cost hospitalization

| Variable | Non-High-Cost Hospitalization | High-Cost Hospitalization | P value |
| --- | --- | --- | --- |
| Age (years, mean +/- SD) | 76.03 +/- 7.98 | 75.89 +/- 7.84 | 0.34 |
| Female (%) | 11,265 (41.3) | 1320 (45.7) | <0.001 |
| White (%) | 84 | 84.8 | 0.2 |
| Charlson Comorbidity Index (median, IQR) | 1[1-3] | 1[1-3] | 0.018 |
| Comorbidities (%) |  |  |  |
| Congestive Heart Failure | 12.6 | 11.2 | 0.001 |
| Coronary Artery Disease | 38.1 | 41.2 | 0.04 |
| Peripheral Vascular Disease | 16.5 | 14.5 | 0.006 |
| Cerebrovascular Disease | 7.6 | 7.3 | 0.56 |
| Chronic Obstructive Pulmonary Disease | 21.9 | 21.6 | 0.72 |
| Chronic Kidney Disease | 24.2 | 20.9 | <0.001 |
| Moderate-Severe Liver Disease | 0.5 | 0.7 | 0.16 |
| Diabetes | 34.8 | 32.9 | 0.035 |
| Income Quartile (%) |  |  |  |
| 76<sup>th</sup>-100<sup>th</sup> | 24.6 | 29.8 | 0.001 |
| 51<sup>st</sup>-75<sup>th</sup> | 28 | 26 | 0.024 |
| 26<sup>th</sup>-50<sup>th</sup> | 25.9 | 24.4 | 0.087 |
| 1<sup>st</sup>-25<sup>th</sup> | 20.0 | 18.2 | 0.023 |
| Payer Status (%) |  |  |  |
| Medicare | 88.9 | 90.1 | 0.083 |
| Medicaid | 1.2 | 0.7 | 0.019 |
| Private | 7.9 | 8.0 | 0.857 |
| Other | 1.9 | 1.2 | 0.011 |
| Hospital Region |  |  | <0.001 |
| New England | 2.5 | 4.5 |  |
| Middle Atlantic | 12.2 | 13.7 |  |
| East North Central | 14.6 | 9.3 |  |
| West North Central | 8.5 | 5 |  |
| South Atlantic | 21.2 | 19.4 |  |
| East South Central | 5.5 | 7.6 |  |
| West South Central | 12 | 13 |  |
| Mountain | 10 | 9.9 |  |
| Pacific | 13.3 | 17.6 |  |
| Hospital Volume |  |  | <0.001 |
| Low Volume (1-15 procedures/year) | 10.6 | 15.9 |  |
| Medium Volume (16-35 procedures/year) | 25.7 | 32.5 |  |
| High Volume (> 35 procedures/year) | 63.7 | 51.6 |  |
| Major Adverse Event (%) | 1060 (3.9) | 325 (11.2) | <0.001 |
| In-Hospital Death | 15 (0.1) | 25 (0.9) | <0.001 |
| Acute MI | 20 (0.1) | - |  |
| Pericardial Effusion/Tamponade requiring pericardiocentesis or surgery | 150 (0.5) | 130 (4.7) | <0.001 |
| Bleeding or Transfusion | 655 (2.4) | 230 (8.0) | <0.001 |
| Stroke (Ischemic/Hemorrhagic) or TIA | 170 (0.6) | <10 (0.2) | 0.002 |
| Systemic Embolization | 25 (0.1) | <10 (0.2) | 0.182 |
| Vascular Complication | 565 (2.1) | 190 (6.6) | <0.001 |
| Median Length of Stay (IQR) | 1 (1) | 1 (1-2) | <0.001 |
| Median Cost (IQR) | \$23,600 (\$18,100-28,400) | \$43,500 (\$39,500-51,300) | <0.001 |

On multivariate analysis, lower procedural volume center, occurrence of MAE, congestive heart failure, high comorbidity burden (CCI =6), and length of stay were independent predictors of a high-cost hospitalization (**Table 3**).

**Table 3.** Multivariate Regression model to predict factors associated with high cost hospitalization.

| Variable | aOR | 95% C.I. | p value |
| --- | --- | --- | --- |
| Age | 0.99 | 0.985-0.996 | 0.001 |
| Female | 1.05 | 0.966-1.142 | 0.25 |
| White | 0.921 | 0.815-1.04 | 0.182 |
| Income Quartile |  |  |  |
| 0-25th | 0.793 | 0.7-0.899 | 0.001 |
| 26-50th | 0.83 | 0.742-0.93 | 0.001 |
| 51-75th | 0.721 | 0.646-0.805 | 0.001 |
| 76-100th | Ref |  |  |
| Payer Status |  |  |  |
| Medicare | 1.309 | 0.922-1.858 | 0.133 |
| Medicaid | 0.373 | 0.195-0.715 | 0.003 |
| Private | 1.119 | 0.767-1.633 | 0.559 |
| Other | Ref |  |  |
| Hospital Region |  |  |  |
| New England | 0.612 | 0.491-0.763 | 0.001 |
| Middle Atlantic | 0.336 | 0.266-0.426 | 0.001 |
| East North Central | 0.374 | 0.288-0.487 | 0.001 |
| West North Central | 0.506 | 0.407-0.629 | 0.001 |
| South Atlantic | 0.9 | 0.702-1.155 | 0.408 |
| East South Central | 0.626 | 0.498-0.786 | 0.001 |
| West South Central | 0.578 | 0.455-0.733 | 0.001 |
| Mountain | 0.811 | 0.653-1.007 | 0.058 |
| Pacific | Ref |  |  |
| Hospital Volume |  |  |  |
| Low Volume Center | 2.11 | 1.875-2.376 | 0.001 |
| Medium Volume Center | 1.694 | 1.544-1.858 | 0.001 |
| High Volume Center | Ref |  |  |
| Major Adverse Event | 1.281 | 1.073-1.53 | 0.006 |
| Comorbidities |  |  |  |
| Coronary Artery Disease | 0.958 | 0.8-1.147 | 0.639 |
| Congestive Heart Failure | 1.25 | 1.072-1.459 | 0.004 |
| Peripheral Vascular Disease | 0.9 | 0.761-1.064 | 0.218 |
| Cerebrovascular Disease | 0.828 | 0.675-1.016 | 0.071 |
| COPD | 0.92 | 0.784-1.079 | 0.305 |
| Diabetes | 0.932 | 0.797-1.091 | 0.384 |
| Chronic Kidney Disease | 0.792 | 0.67-0.936 | 0.006 |
| Moderate-Severe Liver Disease | 0.988 | 0.55-1.774 | 0.966 |
| CCI Score |  |  |  |
| CCI Score =1 | 0.931 | 0.79 -1.098 | 0.396 |
| CCI Score =2 | 0.865 | 0.659-1.135 | 0.295 |
| CCI Score =3 | 0.852 | 0.578-1.256 | 0.418 |
| CCI Score =4 | 0.714 | 0.43-1.188 | 0.195 |
| CCI Score =5 | 0.571 | 0.287-1.138 | 0.111 |
| CCI Score =6 | 2.504 | 1.122-5.59 | 0.025 |
| Length of Stay | 1.412 | 1.369-1.458 | 0.001 |

## Discussion

In this large nationwide study of patients undergoing LAAO with Watchman device, we found significant variation in total hospitalization costs. Nearly 13% of the variability in patient-level costs was related to the center performing the procedure. These differences persisted even after adjustment for patient characteristics (demographics, comorbidities, insurance), procedural complications, hospital location, hospital volume, and length of stay. Similarly, among patients experiencing a major adverse event, substantial variation in costs attributable to the center was observed. Interestingly in our study cohort, less than 1% of the variation in complications was attributable to interhospital differences. Taken together, our findings suggest that differences in hospital care pathways and resource utilization in management of complications, rather than the incidence of procedure-related complications, were major drivers for the observed center-level variation in costs for Watchman device implantation. These interhospital differences in total patient costs were also more prominent for patients undergoing Watchman device implantation at low-volume centers compared to high-volume centers. This could possibly be due to greater operator experience, lower rates of complications, greater availability of services, and utilization of standardized protocols of care at high volume centers^7,8^.

Despite the proliferation of value-based healthcare in the United States, substantial variation in costs across different hospitals among Medicare beneficiaries has been previously reported^9,10^. We observed similar center-level variation in total costs in Medicare beneficiaries receiving a Watchman device. This finding further strengthens our observation that disparities in patient-level costs are, at least in part, due to differences in practice patterns and service use related to individual centers. Amongst non-Medicare patients, such variation was even more pronounced, probably due to the superimposed influence of variation in specific insurer-hospital contracts^11,12^.

We found care at hospitals with low procedural volumes and procedure-related complications independently predictive of high hospital costs. Although our study did not analyze the association between hospital procedural volume and adverse events, an inverse volume-outcome relationship for LAAO procedure has been previously described^6^. This could be due to variation in operator skills and technical proficiency, and management of potential complications at low volume centers, which may lead to increased resource utilization and thus increased costs.

Establishing minimum operator and institutional volume standards for LAAO procedure may potentially result in improved outcomes, thereby reducing unwarranted expenditures^8,13^. Additionally, longer hospital stays were associated with high hospital costs. In contemporary practice, patients are hospitalized overnight after LAAO and typically discharged the following day. Same day discharges have begun to occur but are not widespread. With increasing experience and improving outcomes, same-day discharge in selected patients could be feasible, in doing so reducing length of stay and overall costs^14^.

As the commercial use of percutaneous LAAO with the Watchman device increases in the United States, the wide variability of Watchman-related costs underscores the importance of standardizing hospital practices to achieve high-quality value-based care. Previous cost-effectiveness analyses of the Watchman device have suggested it to be an economically viable stroke risk reduction strategy^4,5^. However, such analyses are highly dependent on input variables. The median hospitalization cost in our study was substantially higher than the cost inputs used in prior analyses ($24,500 vs. $16,800), suggesting Watchman implantation could be less cost-effective than previously thought. Therefore, addressing unwarranted cost variation could improve the cost-effectiveness of Watchman device, further expanding the economic appropriateness of this procedure to a wider population.

## Limitations

To our knowledge, this is the first study to explore center level differences in cost for LAAO with Watchman device. In this analysis, we used a large administrative database to evaluate costs. A strength of the present analysis was the use of cost data rather than charges to better reflect the cost of the services being provided. These data probably are an underestimate of true total costs as they do not account for physician professional fees, and other costs relevant to the family and society such as transportation to the hospital, and loss of income and productivity if it is necessary to take time off from the workplace. The NIS Database does not track hospitals across years, thereby preventing examination of changes in cost variation at individual hospitals over the study period. Although administrative data sources contain valuable resource utilization information, they lack data on procedural details, medication regimens, costs associated with specific phases of care (Watchman device price, operating room time, anesthesia, intensive care unit stay, etc.). It is possible that certain unmeasured confounders may be present; however, we were able to adjust for important factors demonstrated to affect resource utilization. It is possible that coding errors may exist in administrative data sources.

## Conclusion

In this national-level analysis of hospital costs in patients undergoing LAAO with Watchman device, we found significant variation from center to center even after accounting for important patient factors, hospital volume and location, and length of stay. As the burden of AF increases and greater number of institutions utilize this therapy, examination of factors associated with hospital-based variation is warranted as to develop value-based healthcare systems.

## Data Availability

The data described in this article are available at https://hcup-us.ahrq.gov

## Acknowledgements

None

